# Aquaporin-4 polymorphisms modify the penetrance of Parkinson’s disease in *leucine-rich repeat kinase 2* carriers

**DOI:** 10.1101/2025.10.04.25337325

**Authors:** Mariateresa Buongiorno, Natalia Vilor-Tejedor, Bárbara Segura, Alex Iranzo, Yaroslau Compta, Clara Marzal-Espí, Darly Milena Giraldo, Jorge Hernández-Vara, Victoria González, Oriol de Fábregues, Pilar Delgado, Jerzy Krupinski, Oriol Grau-Rivera, Gonzalo Sánchez-Benavides

## Abstract

**Background:** Pathogenic mutations in the *leucine-rich repeat kinase-2* (*LRRK2*) gene are the most common cause of familial Parkinson’s disease (PD). *LRRK2* shows incomplete penetrance, yet the biological factors influencing disease expression remain unknown.

**Objectives:** To investigate whether genetic variants in aquaporin-4 (*AQP4*), key in glymphatic system functioning, are associated with the penetrance of PD in *LRRK2* carriers.

**Methods:** We analyzed baseline data from 302 *LRRK2* carriers from the Parkinson’s Progression Markers Initiative. Fourteen *AQP4* single nucleotide polymorphisms, previously implicated in PD, Alzheimer’s disease, or other neurodegenerative-related processes were tested for association with PD manifestation using logistic regression models adjusted for age and sex. Recessive, dominant, and additive genetic models were explored. Sensitivity analyses were conducted in G2019S carriers (n=273).

**Results:** One hundred twenty-seven (42%) *LRRK2* carriers were asymptomatic, and 174 (58%) had PD. There were no differences between groups in age (63.5[9.5] vs. 62.2[7.5]) or number of women (52.0% vs. 55.7%). Homozygosity for the minor allele of rs9951307 was associated with reduced likelihood of PD (OR=0.28, 95% CI 0.10–0.64, p=0.005), whereas rs335930 homozygosity was associated with increased likelihood (OR=4.2, 95% CI 1.41–15.6, p=0.016). Additive models supported these associations, though rs335930 did not surpass the adjusted threshold. Results were consistent in the G2019S subgroup.

**Conclusions:** *AQP4* polymorphisms may contribute to the variable penetrance of *LRRK*2 mutations, potentially though modulation of glymphatic clearance. These findings support the glymphatic system as a relevant pathway in familial PD and highlight *AQP4* as a candidate therapeutic target.

## Introduction

Parkinson’s disease (PD) is the fastest growing neurodegenerative disease (1), clinically characterized by cardinal motor features (bradykinesia, rigidity, resting tremor, and postural instability), non-motor symptoms, and pathologically by dopaminergic neuronal loss and alpha-synuclein-containing Lewy-type pathology (2). Pathogenic mutations in the *leucine-rich repeat kinase-2* (*LRRK2*) gene are the most frequent known genetic cause of familial and some sporadic PD (1% of the PD), with the G2019S substitution being the most prevalent variant worldwide (3–6). Despite sharing clinical features with idiopathic PD (7), LRRK2-associated PD displays incomplete and variable penetrance, ranging from 25% in non-Ashkenazi populations to more than 90% in North Africans by age 80 (8,9). The reasons why some carriers develop PD while others remain unaffected are not fully understood.

A growing body of work has explored potential genetic and environmental modifiers of penetrance. Genome-wide and candidate studies have implicated loci such as *CORO1C* (10), *DNM3* (11), *SNCA* and *MAPT* (12,13), as well as mitochondrial DNA abnormalities (14–16) in manifesting G2019S carriers, and polygenic risk scores (17,18). Environmental exposures including smoking, coffee intake, and nonsteroidal anti-inflammatory drug use appear protective, whereas pesticides increase risk (19,20). Yet, the effect sizes of individual modifiers remain modest, and the major drivers of penetrance in LRRK2 carriers are still unknown (21).

One candidate pathway that has gained recent attention is the glymphatic system, a cerebrospinal fluid-based clearance mechanism that facilitates the removal of interstitial waste and proteins through perivascular astrocytic aquaporin-4 (*AQP4*) water channels (22,23). Glymphatic clearance is most active during slow-wave sleep (24) and has been implicated in the accumulation of pathogenic proteins across neurodegenerative diseases (25). Imaging studies show reduced glymphatic function in idiopathic PD, correlating with worse clinical outcomes (26–29), and has been also observed in isolated REM sleep behaviour disorder (iRBD), a prodromal synucleinopathy stage (30). Importantly, recent work suggests altered glymphatic function also in *LRRK2* carriers, with larger perivascular space volumes in manifesting versus non-manifesting individuals (31,32).

To date, only two studies have focused on glymphatic dysfunction in *LRRK2* carriers. Donahue et al., using Parkinson’s Progression Markers Initiative (PPMI) data, compared perivascular space (PVS) fraction volume as a glymphatic function proxy among controls, idiopathic PD and manifesting and non-manifesting carriers of pathogenic mutations (*LRRK2, SNCA* and *GBA*) and found that PVS volume fraction was higher in symptomatic PD, both idiopathic and familiar, but was driven mainly by a marked increase between manifesting and non-manifesting *LRRK2* carriers (32). More recently, Zhang et al., analyzed longitudinal data from non-manifesting *LRRK2* and *GBA* carriers and likewise observed lower glymphatic function, as measured by DTI-along perivascular spaces (ALPS) index, in non-manifesting carriers than healthy controls. They also found that baseline DTI-ALPS predicted cognitive decline, and that increasing ALPS index was associated with a reduced risk of disease progression (31).

It is known that glymphatic function is affected by genetic variation in AQP4. Specific *AQP4* single nucleotide polymorphisms (SNPs) have been linked to amyloid-β deposition in Alzheimer’s disease (AD) (33,34), risk of PD (35), and cognitive outcomes in PD (36) and AD (34,37). Moreover, experimental evidence indicates a direct molecular interaction between *LRRK2* and *AQP4*: *LRRK2*-mediated phosphorylation disrupts AQP4 perivascular localization, impairing glymphatic clearance and exacerbating neuroinflammation and dopaminergic neurodegeneration in animal models (38). These findings led us to hypothesize a biologically plausible role for *AQP4* genetic variation in modifying *LRRK2* penetrance.

Here, to test this hypothesis we use PPMI data to investigate whether common *AQP4* polymorphisms are associated with clinically manifest PD among *LRRK2* mutation carriers. We focused on 14 SNPs previously implicated in PD, AD, or sleep-related processes, and applied logistic regression models under recessive, dominant, and additive frameworks, adjusting for age and sex. Given the predominance of the G2019S variant in PPMI, sensitivity analyses were restricted to this subgroup. We hypothesized that *AQP4* variants influencing glymphatic function would be associated with differences in *LRRK2* penetrance.

## Methods

### Participants

We selected the 406 carriers of the pathogenic *LRRK2* mutation enrolled in the Parkinson’s Progression Markers Initiative (PPMI). Participants were classified at study entry as manifesting PD (mPD) or non-manifesting PD (non-mPD) based on clinical diagnosis made by site investigator per established diagnostic criteria with confirmatory DAT-SPECT required. Only participants with valid genotyped data to impute *AQP4* SNPs (n=302) were included in the analysis. Analyzed participants were enrolled between September 2013 and January 2022 in PPMI protocol was approved by the institutional review board of each participating site in PPMI, and all participants gave written informed consent. PPMI information, protocol and manuals are available at ppmi- info.org/studydesign.

### Selection of genetic variants, and imputation

We obtained genotyped data from PPMI, generated using Illumina ImmunoChip or NeuroX arrays.Imputation was conducted using the Michigan Imputation Server, with the Haplotype Reference Consortium, reference panel (GRCh38). From the imputed dataset, we extracted 14 SNPs within the *AQP4* gene previously reported as relevant in PD, AD, or related neurodegenerative processes (see Table 1 for details and Supplementary Materials for search strategy and terms used).

**Table 1.**
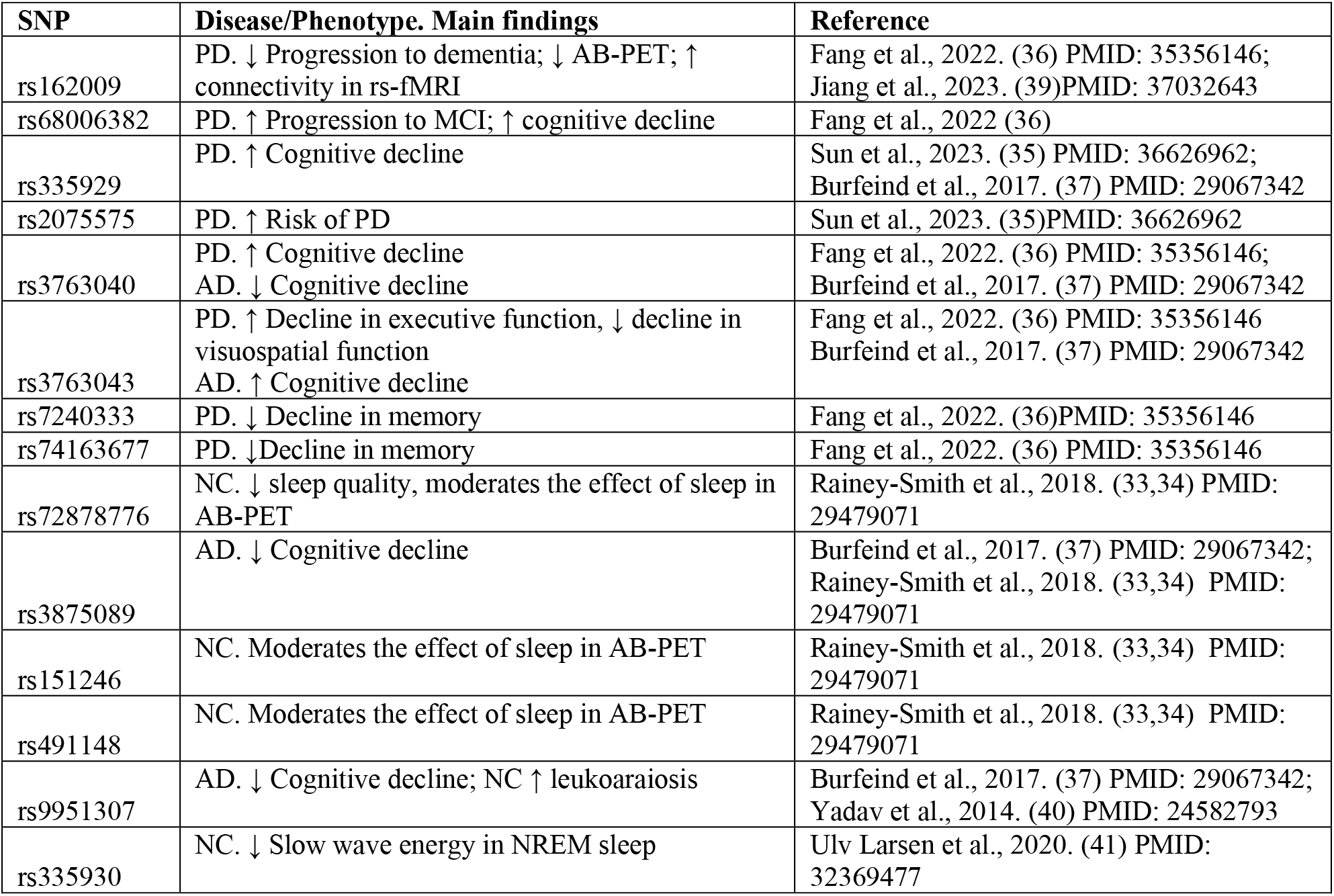
AQP4 SNPs selected from the literature.

### Statistical Analysis

We summarized continuous variables as means and standard deviations, and categorical variables as count and percentages. We compared demographics and clinical characteristics between manifesting PD (mPD) and non-manifesting PD (non-mPD) carriers using two-sample t-tests and chi-square tests as appropriate We assessed the association between *AQP4* variants and PD manifestation using logistic regression. We modeled PD status (manifesting vs non-manifesting) as the dependent variable, included each SNP as the predictor of interest, and adjusted for age and sex as covariates. As *LRRK2* G2019S variant was the most common one in the sample (90.4%) a sensitivity analyses restricted to G2019S carriers was performed. We examined three genetic inheritance assumptions: in the recessive model we contrasted homozygous minor allele carriers against all others; in the dominant model we contrasted carriers of at least one minor allele against homozygous major allele carriers; and in the additive model we coded genotypes as 0, 1, or 2 to test for a linear trend in the log-odds of disease. We estimated odds ratios (ORs) and 95% confidence intervals (CIs) by exponentiating regression coefficients, and we derived p-values from Wald tests. Recessive models were restricted to those SNPs that showed at least 5% frequency of the 2 minor alleles in the sample (rs162009, rs68006382, rs2075575, rs3763043, rs9951307, rs335930).

To account for non-independence among the 14 SNPs due to linkage disequilibrium (LD), we grouped them into clusters based on pairwise LD estimates. We computed both the squared correlation coefficient (r^2^) and normalized LD measure (D′) using LDmatrix from the LDlink suite (v5.4), referencing GRCh38 High Coverage data from 1000 Genomes Phase 3. Several SNP pairs showed high LD (D′ or r^2^ > 0.8), so we used hierarchical clustering with Ward’s method on a dissimilarity matric (1-D’) to define LD blocks. This revealed three distinct clusters consistent with the LD heatmap (Supplemental Figure 1), which were treated as approximately independent. For multiple testing correction, we used the number of LD clusters as a proxy for the effective number of independent tests setting the statistical significance threshold at p<0.017 (0.05/3). This approach balances the conservativeness of standard Bonferroni correction and the risk of underestimating false positives by ignoring LD structure.

## Results

### Demographic and clinical data

From the 302 *LRRK2* variant carriers included in the study, 127 (42%) were mPD and 175 (58%) were non-mPD. The mean age of mPD individuals was slightly but not significantly higher than that of non mPD (63.5 y.o. vs 62.3 y.o., p=0.22). Sex distribution was similar between groups (mPD 52% vs non-mPD 56% of women, p=0.56). The most prevalent pathogenic variant was G2019S (90.4%), followed by R1441G (8.9%), with minimal representation of R1414C (0.3%) and N1437H (0.3%), see Table 2.

**Table 2.**
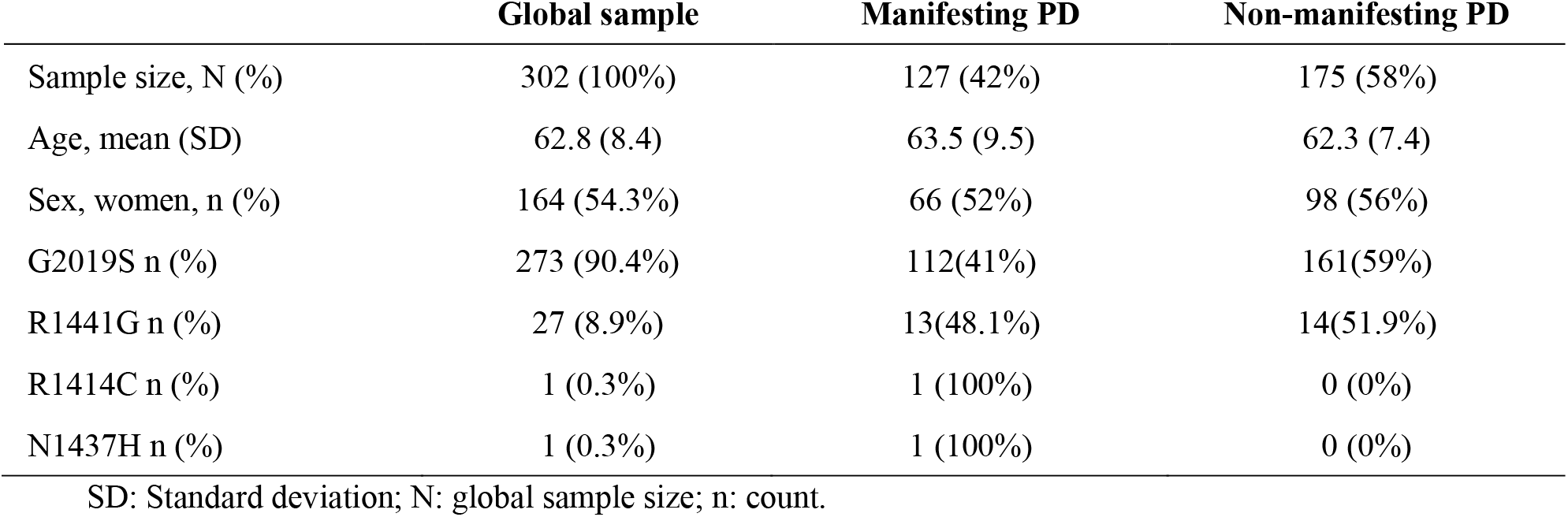
Clinical and demographic characteristics.

|  | Global sample | Manifesting PD | Non-manifesting PD |
| --- | --- | --- | --- |
| Sample size, N (%) | 302 (100%) | 127 (42%) | 175 (58%) |
| Age, mean (SD) | 62.8 (8.4) | 63.5 (9.5) | 62.3 (7.4) |
| Sex, women, n (%) | 164 (54.3%) | 66 (52%) | 98 (56%) |
| G2019S n (%) | 273 (90.4%) | 112(41%) | 161(59%) |
| R1441G n (%) | 27 (8.9%) | 13(48.1%) | 14(51.9%) |
| R1414C n (%) | 1 (0.3%) | 1 (100%) | 0 (0%) |
| N1437H n (%) | 1 (0.3%) | 1 (100%) | 0 (0%) |

SD: Standard deviation; N: global sample size; n: count.

### AQP4 SNPs prediction of PD manifestation

Under recessive genetic models (homozygous minor allele vs. heterozygous/major allele), rs9951307 showed a protective association with mPD (OR = 0.27 CI 95% [0.10- 0.64]; p = 0.005), while rs335930 was associated with increased risk (OR = 4.23; CI 95% [1.41-15.6]; p = 0.016) (Table 3, Figure 1). Dominant models (carriers of ≥ 1 minor allele vs non-carriers) revealed no significant associations.

**Table 3.** Results from the logistic regressions for recessive (0/1 vs 2 minor alleles), dominant (0 vs 1/2 minor alleles) and additive (being carrier of 0, 1 or 2 of the minor allele) models in the prediction of presence of PD in *LRRK2* carriers adjusted by age and sex. LD: Linkage disequilibrium; SNP: single nucleotid polymorphism; OR: Odds ratio; *n=296; -: non- computed due to frequency of the minor allele less than 5%. Significant associations (p<0.017) are shown in bold.

| LD cluster | SNP | Minor allele | Recessive model |  | Dominant model |  | Additive model |  |
| --- | --- | --- | --- | --- | --- | --- | --- | --- |
|  |  |  | OR [95% CI] | p-value | OR [95% CI] | p-value | OR [95% CI] | p-value |
| 1 | rs162009 | A | 1.36 [0.67–2.72] | 0.387 | 1.48 [0.92–2.39] | 0.102 | 1.32 [0.94–1.86] | 0.109 |
| 1 | rs68006382* | G | 1.74 [0.72–4.28] | 0.217 | 0.87 [0.54–1.43] | 0.624 | 1.03 [0.71–1.48] | 0.888 |
| 1 | rs335929* | C | - | - | 1.49 [0.93–2.40] | 0.100 | 1.62 [1.08–2.43] | 0.020 |
| 1 | rs335930* | C | **4.23 [1.41–15.6]** | **0.016** | 1.46 [0.91–2.34] | 0.116 | 1.58 [1.07–2.35] | 0.023 |
| 1 | rs3763043 | T | 1.16 [0.54–2.47] | 0.703 | 1.10 [0.70–1.76] | 0.673 | 1.09 [0.77–1.55] | 0.622 |
| 2 | rs7240333* | T | - | - | 1.08 [0.57–2.03] | 0.816 | 1.04 [0.58–1.86] | 0.885 |
| 2 | rs74163677* | A | - | - | 0.46 [0.18–1.03] | 0.070 | 0.45 [0.19–1.00] | 0.062 |
| 2 | rs72878776* | A | - | - | 0.86 [0.49–1.48] | 0.587 | 0.83 [0.50–1.36] | 0.460 |
| 2 | rs3763040* | A | - | - | 1.35 [0.82–2.23] | 0.236 | 1.28 [0.81–2.03] | 0.285 |
| 2 | rs9951307 | G | **0.27 [0.10–0.64]** | **0.005** | 0.78 [0.49–1.24] | 0.300 | 0.69 [0.48–0.97] | 0.036 |
| 3 | rs3875089 | C | - | - | 0.82 [0.49–1.36] | 0.438 | 0.84 [0.53–1.31] | 0.450 |
| 3 | rs151246* | T | - | - | 1.18 [0.71–1.98] | 0.515 | 1.21 [0.76–1.93] | 0.415 |
| 3 | rs491148 | G | - | - | 0.76 [0.45–1.25] | 0.282 | 0.89 [0.58–1.33] | 0.560 |
| 3 | rs2075575* | A | 1.05 [0.58–1.88] | 0.878 | 1.16 [0.71–1.93] | 0.558 | 1.08 [0.78–1.50] | 0.637 |

**Figure 1.**
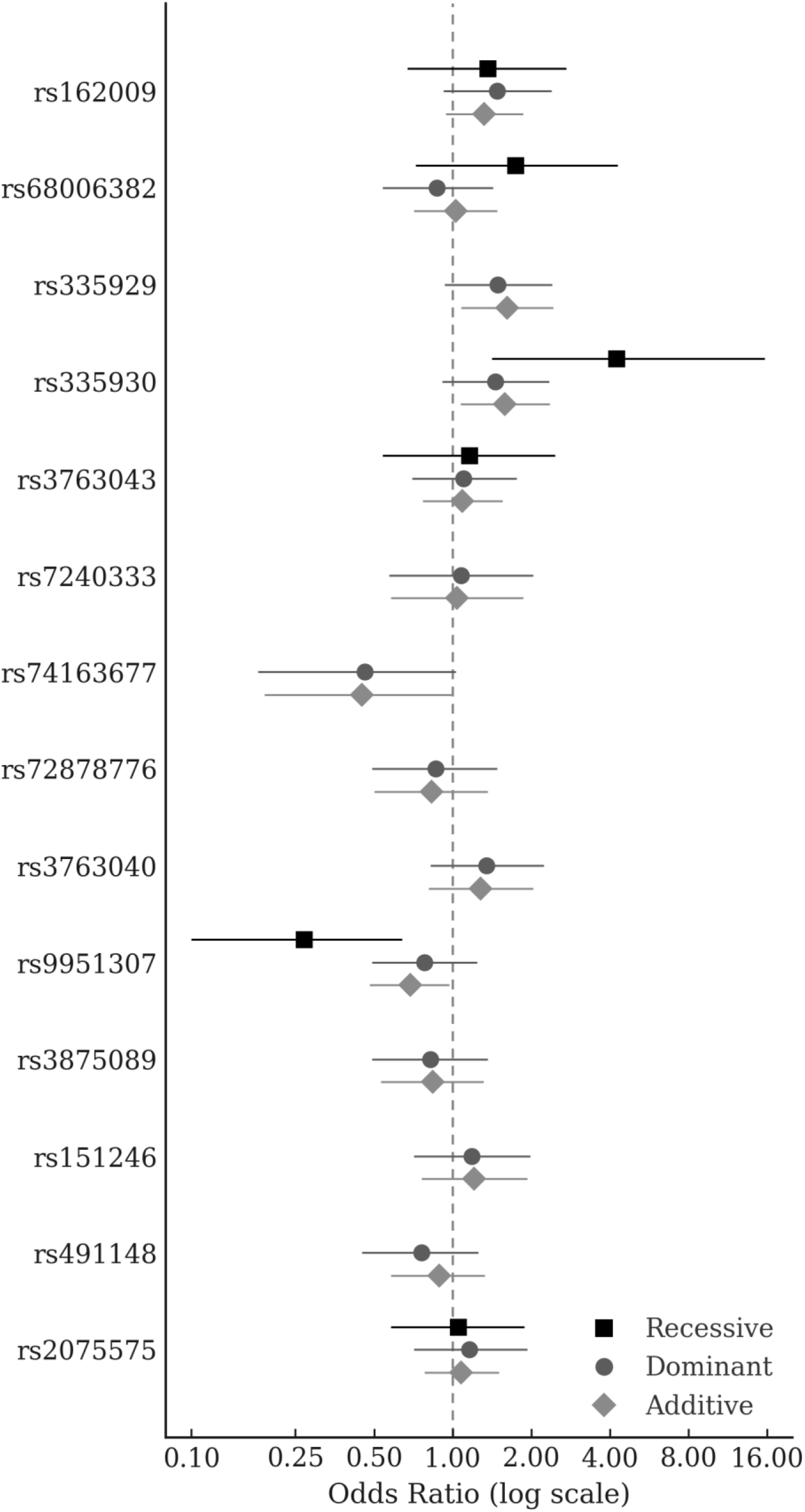
Odds Ratios and 95% CI for recessive (0/1 vs 2 minor alleles), dominant (0 vs 1/2 minor alleles) and additive (being carrier of 0, 1 or 2 minor alleles) models in the prediction of presence of PD in *LRRK2* carriers adjusted by age and sex.

In additive models (linear increase on effect per minor allele), results were consistent with the recessive findings. rs9951307 remained protective (OR = 0.69; CI95% [0.48-0.97]; p = 0.036), whereas rs335929 (OR = 1.62; CI95% [1.08-2.43]; p = 0.020) and rs335930 (OR = 1.58; CI95% [1.07-2.35]; p = 0.023) were associated with increased risk. Despite these trends, the observed p-value did not reach the adjusted significance threshold (p=0.017).

### Sensitivity analyses

Analyses restricted to G2019S variant carriers (n=273) yielded results consistent with those of the full cohort, with comparable effect sizes and p-values (See Supplementary Table S1, and Figure S2).

## Discussion

In this study, we cross-sectionally explored the association between *AQP4* SNPs and PD manifestation in *LRKK2* mutation carriers. To our knowledge, this is the first investigation adding evidence suggesting that the genetic variability of *AQP4*, a key element in glymphatic function, may help explain incomplete penetrance in familial PD.

We found that homozygosity for the minor allele (G) of rs9951307 was associated with lower penetrance, reducing the likelihood of manifesting PD in 73%, while homozygosity for the minor allele (C) of rs335930 substantially increased risk of disease expression.

*AQP4* is encoded by a 3-kb gene on chromosome 18. *AQP4* encodes the principal water channel of astrocytic endfeet and plays a central role in glymphatic clearance, astrocytic polarity, and neuroinflammatory signaling. The use of two alternative transcription start sites generates the M1 and M23 isoforms, whose distinct N-terminal features determine their ability to assemble into orthogonal arrays, thereby modulating membrane organization and water permeability (42,43).

Previous work has implicated *AQP4* variants in sporadic PD, such as the association of rs2075575 reported in Han Chinese (35), although we did not replicate this finding in the context of *LRRK2*-related disease. Such discrepancies may reflect differences in pathophysiology between sporadic and familial PD, population ancestry, or specific interactions between *LRRK2* and *AQP4* (38).

The protective effect of rs9951307 aligns with findings in AD, where minor allele carriers showed slower cognitive decline and fewer vascular lesions (37). By contrast, Liu et al. recently reported less favorable glymphatic-related changes in cognitively unimpaired older adults with the same variant (44), highlighting that its effects may vary across clinical contexts. For rs335930, our results are consistent with prior evidence linking this polymorphism to altered slow-wave sleep and glymphatic efficiency (41), as well as increased amyloid burden in risk-score analyses (45). Together, these observations suggest that *AQP4* variation influences penetrance of *LRRK2* mutations through pathways involving astrocytic water transport and protein clearance

The magnitude of the penetrance effects found in the present study were larger than those seen in polygenic risk score analyses of *LRRK2* G2019S carriers (OR=1.28 per 1 standard deviation (18) and OR=1.49 (17), but lower than those reported in recent genome-wide analyses (10).Our findings suggest the contribution of another physiological pathway in which the variability in glymphatic function, modulated by *AQP4* genetic polymorphisms that tightly interact with *LRRK2*, plays a key role in the development of PD.

Experimental evidence supports this interpretation. In a recent study Lopes and colleagues (46) used a mouse model of α-synuclein propagation to examine how glymphatic function modulates its spread, and how propagating α-synuclein affects glymphatic dynamics in turn. After intrastriatal injection of α-synuclein pre-formed fibrils, they observed a marked reduction in the expression of *AQP4* in astrocytic that inversely correlated with phosphorylated α-synuclein burden. As pathology progressed to the midbrain, glymphatic transport increased, seemingly accommodating the incoming α- synuclein and partially normalizing regional protein load. To study causality, the authors pharmacologically inhibited glymphatic clearance and observed that acute inhibition reduced α-synuclein efflux from brain to CSF, and sustained inhibition in seeded mice aggravated striatal α-synuclein pathology, produced striatal and midbrain atrophy, and worsened behavioral performance. The same group has already demonstrated that the pharmacological inhibitor of AQP4, TGN-020, increased tau aggregation and propagation in transgenic mice, aggravating memory performance (47). Thus, glymphatic dysfunction would also affect the clearance of other pathological proteins and neuropathological examinations of *LRRK2* carriers brains consistently found neurofibrillary tangles, neuritic plaques and neuropil threads (48). These mechanistic and neuropathological findings, as well as the observation that pathogenic *LRRK2* variants increase *AQP4* phosphorylation, leading to loss of perivascular polarity (38), lend support to our hypothesis that *AQP4* genetic variability may impact the penetrance of *LRRK2* by modulating glymphatic clearance

From a clinical perspective, these results suggest that *AQP4* genotype might contribute to stratification strategies for *LRRK2* carriers. Integrating *AQP4* variants into predictive models could improve identification of individuals at higher risk for conversion. In parallel, imaging and fluid biomarkers of glymphatic function could provide useful intermediate measures for future studies. More broadly, interventions aimed at enhancing glymphatic clearance, either pharmacologically or through lifestyle approaches, may represent promising strategies to delay disease onset in genetically at-risk groups.

The present results open new avenues of research. As next steps we will explore the association between glymphatic function, *AQP4* genetic variability, and positivity or negativity of aSyn SAA/RT-QuIC, since it is unknown why some familial forms of PD only have neurodegeneration in substantia nigra pars compacta in absence of Lewy bodies, while the majority display the full neuropathological expression.

This study is not free of limitations. Although we assessed a comprehensive set of *AQP4* SNPs, it is possible that other polymorphisms not included in our analysis have a role in the penetrance of *LRRK2*. Despite correction for multiple testing within LD clusters and models, type I errors cannot be fully excluded. The cross-sectional nature of the study limited the ability to test longitudinal outcomes and age of onset related effects of *AQP4* SNPs. Finally, the findings of this study cannot be directly applicable in other populations, such Asian, in which G2019S variant is rare or absent (49).

In summary, our findings support that *AQP4* genetic variants may play a role in the penetrance of *LRRK2* pathogenic mutations, supporting the influence of glymphatic function in the pathogenesis of PD in familial PD. Replication in larger and multi-ancestry cohorts, ideally with longitudinal follow-up, will be necessary to confirm these results. If validated, targeting glymphatic pathways could open new preventive pharmacological approaches for familial PD.

## Acknowledgements

Data used in the preparation of this article was obtained on the 1^st^ of April of 2024 from the Parkinson’s Progression Markers Initiative (PPMI) database (www.ppmi-info.org/access-dataspecimens/download-data), RRID:SCR_006431. For up-to-date information on the study, visit www.ppmi-info.org. PPMI -a public-private partnership- is foundered by The Michael J. Fox Foundation for Parkinson’s Research and funding partners, including AbbVie, AcureX Therapeutics, Allergan, Amathus Therapeutics, Avid Radiopharmaceuticals, Bial Biotech, Biogen, BioLegend, Bristol Myers Squibb, Celgene, Dacapo Brainscience, Denali, GE Healthcare, Genentech, GlaxoSmithKline, Lilly, Lundbeck, Merck, Meso Scale Diagnostics, Neurocrine Biosciences, Pfizer, Piramal Imaging, Prevail Therapeutics, Roche, Sanofi Genzyme, Servier, Takeda, Teva, UCB, Vanqua Bio, Verily Life Science, Voyager Therapeutics, and Golub Capital.

NV-T is supported by William H. Gates Sr. Fellowships from the Alzheimer’s Disease Data Initiative, the Spanish Ministry of Science and Innovation - State Research Agency grant RYC2022-038136-I, cofunded by the European Union FSE+, grant PID2022- 143106OA-I00 cofunded by the European Union FEDER, grant 23S06083-001 funded by the Ajuntament de Barcelona and “la Caixa” Foundation, and grant WE.08-2024-07 funded by AlzheimerNederland. BS was supported by Spanish Ministry of Economy and Competitiveness PID2023146932NB-I00 funded by MCI U/AEI/110.13039/5011000111033/FEDER, UE Generalitat de Catalunya (SGR 2021SGR00801) and by María de Maeztu Unit of Excellence (Institute of Neurosciences, University of Barcelona) CEX2021-001159-M, the Spanish Ministry of Science and Innovation. GS-B receives support from the grant CP23/00039, funded by the Instituto de Salud Carlos III (ISCIII) and co-funded by the European Union/FSE+ and the MCIN/AEI/10.13039/501100011033, through the project PID2020-119556RA-I00. OGR receives funding from the Alzheimer’s Association Research Fellowship Program (2019- AARF-644568), from Instituto de Salud Carlos III (ISCIII) through the projects “PI19/00117” and “PI24/00116”, both co-funded by the European Union/FEDER, and from the grant IJC2020-043417-I, funded by MCIN/AEI/10.13039/501100011033 and the European Union NextGenerationEU/PRTR.

## Authors’ Roles

MB (design, execution, interpretation of the data, writing, editing of final version of the manuscript), NV-T (design, execution, analysis, interpretation of the data, writing, editing of final version of the manuscript), BS (interpretation of the data, critical review, editing of final version of the manuscript), AI (critical review, editing of final version of the manuscript), YC (critical review, editing of final version of the manuscript), CM-E (critical review, editing of final version of the manuscript), DM-G (critical review, editing of final version of the manuscript), JH-V (critical review, editing of final version of the manuscript), VG (critical review, editing of final version of the manuscript), OdF (critical review, editing of final version of the manuscript), PD (critical review, editing of final version of the manuscript), JK (critical review, editing of final version of the manuscript), OG (critical review, editing of final version of the manuscript), GS-B (design, execution, analysis, writing, editing of final version of the manuscript).

## Financial Disclosures for the previous 12 months

MB has given paid lectures for Zambon, Bial, and Orion Pharma. GS-B has travel grants from the Alzheimer’s Association. O.G.R. receives research funding from F. Hoffmann‐La Roche Ltd. and has given lectures at symposia sponsored by Roche Diagnostics, S.L.U.

## Data Availability Statement

Data used in the preparation of this article were obtained from the Parkinson Progression Markers Initiative database (ppmi-info.org/access-data-specimens/downloaddata). For up-to-date information on the study, visit ppmi-info.org. GS-B MB, and NT-V takes responsibility for the integrity of the data and the accuracy of the data analysis.

## Notes

### Competing Interest Statement

The authors have declared no competing interest.

### Funding Statement

This study did not receive any funding. PPMI -a public-private partnership- is foundered by The Michael J. Fox Foundation for Parkinson Research and funding partners, including AbbVie, AcureX Therapeutics, Allergan, Amathus Therapeutics, Avid Radiopharmaceuticals, Bial Biotech, Biogen, BioLegend, Bristol Myers Squibb, Celgene, Dacapo Brainscience, Denali, GE Healthcare, Genentech, GlaxoSmithKline, Lilly, Lundbeck, Merck, Meso Scale Diagnostics, Neurocrine Biosciences, Pfizer, Piramal Imaging, Prevail Therapeutics, Roche, Sanofi Genzyme, Servier, Takeda, Teva, UCB, Vanqua Bio, Verily Life Science, Voyager Therapeutics, and Golub Capital.

### Author Declarations

This study was conducting using Parkinson's Progression Markers Initiative (PPMI). PPMI possesses all the necessary ethical approvals from IRBs.

