## Supplementary Materials for "Aquaporin-4 polymorphisms modify the penetrance of Parkinson’s disease in *leucine-rich repeat kinase 2* carriers"

**SUPPLEMENTARY MATERIAL**

**PubMed literature search strategy and terms used to select AQP4 SNPs of interest.**

Search was performed in January 2024 through the PubMed web portal (<https://pubmed.ncbi.nlm.nih.gov/>). Terms used to retrieve the relevant publications were the following:

((AQP4[tiab] OR "aquaporin 4"[tiab] OR "aquaporin-4"[tiab])

AND

(polymorphism*[tiab] OR "single nucleotide polymorphism"[tiab] OR "single nucleotide polymorphisms"[tiab] OR SNP[tiab] OR SNPs[tiab] OR "genetic variant*"[tiab] OR variant*[tiab] OR allele*[tiab] OR haplotype*[tiab] OR genotype*[tiab] OR "rs"[tiab] OR GWAS[tiab] OR "genome wide association"[tiab] OR "whole exome"[tiab] OR sequencing[tiab]))

AND

(Alzheimer*[tiab] OR Parkinson*[tiab] OR dementia*[tiab] OR "Lewy bod*"[tiab] OR DLB[tiab] OR neurodegenerat*[tiab] OR "alpha-synuclein"[tiab] OR "amyloid"[tiab] OR tau[tiab] OR LRRK2[tiab] OR "leucine-rich repeat kinase 2"[tiab] OR GBA[tiab] OR "glucocerebrosidase"[tiab] OR GBA1[tiab]) AND (phenotype*[tiab] OR risk*[tiab] OR susceptib*[tiab] OR severity[tiab] OR progression[tiab] OR survival[tiab] OR outcome*[tiab] OR prognosis[tiab] OR biomarker*[tiab] OR cognition[tiab] OR memory[tiab] OR sleep[tiab] OR clearance[tiab] OR CSF[tiab] OR imaging[tiab] OR MRI[tiab] OR PET[tiab]))

**Table S1.** Results from the logistic regressions for recessive (0/1 vs 2 minor alleles), dominant (0 vs 1/2 minor alleles) and additive (being carrier of 0, 1 or 2 of the minor allele) models in the prediction of presence of PD in G2019S carriers (n=273) adjusted by age and sex.

| **LD cluster** | **SNP** | **Minor allele** | **Recessive model** | | **Dominant model** | | **Additive model** | |
| --- | --- | --- | --- | --- | --- | --- | --- | --- |
|  |  |  | **OR [95% CI]** | **p-value** | **OR [95% CI]** | **p-value** | **OR [95% CI]** | **p-value** |
| 1 | rs162009 | A | 1.51 [0.75−3.06] | 0.250 | 1.51 [0.92−2.52] | 0.109 | 1.39 [0.97−1.99] | 0.071 |
| 1 | rs68006382* | G | 1.92 [0.73−5.20] | 0.188 | 0.89 [0.53−1.49] | 0.666 | 1.04 [0.70−1.55] | 0.837 |
| 1 | rs335929* | C | - | - | 1.55 [0.88−2.41] | 0.147 | 1.61 [1.06−2.47] | 0.026 |
| 1 | rs335930* | C | **4.54 [1.41-16.6]** | **0.011** | 1.43 [0.86-2.36] | 0.164 | 1.58 [1.05-2.40] | 0.029 |
| 1 | rs3763043 | T | 1.17 [0.51−2.62] | 0.700 | 1.00 [0.61−1.64] | 0.996 | 1.03 [0.71−1.50] | 0.858 |
| 2 | rs7240333* | T | - | - | 0.95 [0.48−1.84] | 0.882 | 0.94 [0.50−1.71] | 0.832 |
| 2 | rs74163677* | A | - | - | 0.44 [0.17−1.03] | 0.073 | 0.44 [0.17−0.99] | 0.064 |
| 2 | rs72878776* | A | - | - | 0.99 [0.55−1.74] | 0.965 | 0.93 [0.55−1.55] | 0.776 |
| 2 | rs3763040* | A | - | - | 1.30 [0.77−2.19] | 0.328 | 1.24 [0.77−2.01] | 0.370 |
| 2 | rs9951307 | G | **0.22 [0.06-0.59]** | **0.006** | 0.76 [0.47-1.25] | 0.286 | 0.66 [0.45-0.96] | 0.034 |
| 3 | rs3875089 | C | - | - | 0.91 [0.53−1.45] | 0.731 | 0.92 [0.57−1.47] | 0.738 |
| 3 | rs151246* | T | - | - | 1.21 [0.70−2.06] | 0.494 | 1.19 [0.73−1.96] | 0.726 |
| 3 | rs491148 | G | - | - | 0.77 [0.45−1.31] | 0.282 | 0.94 [0.60−1.46] | 0.792 |
| 3 | rs2075575* | A | 1.06 [0.56−1.98] | 0.856 | 1.09 [0.64−1.85] | 0.338 | 1.06 [0.74−1.50] | 0.744 |

LD: Linkage disequilibrium; SNP: single nucleotid polymorphism; OR: Odds ratio; *n=266; -: non-computed due to frequency of the minor allele less than 5%. Significant associations (p<0.017) are shown in bold.

**Figure S1.** AQP4 SNPs Linkage Disequilibrium (LD) clustering based on pairwise LD estimates.

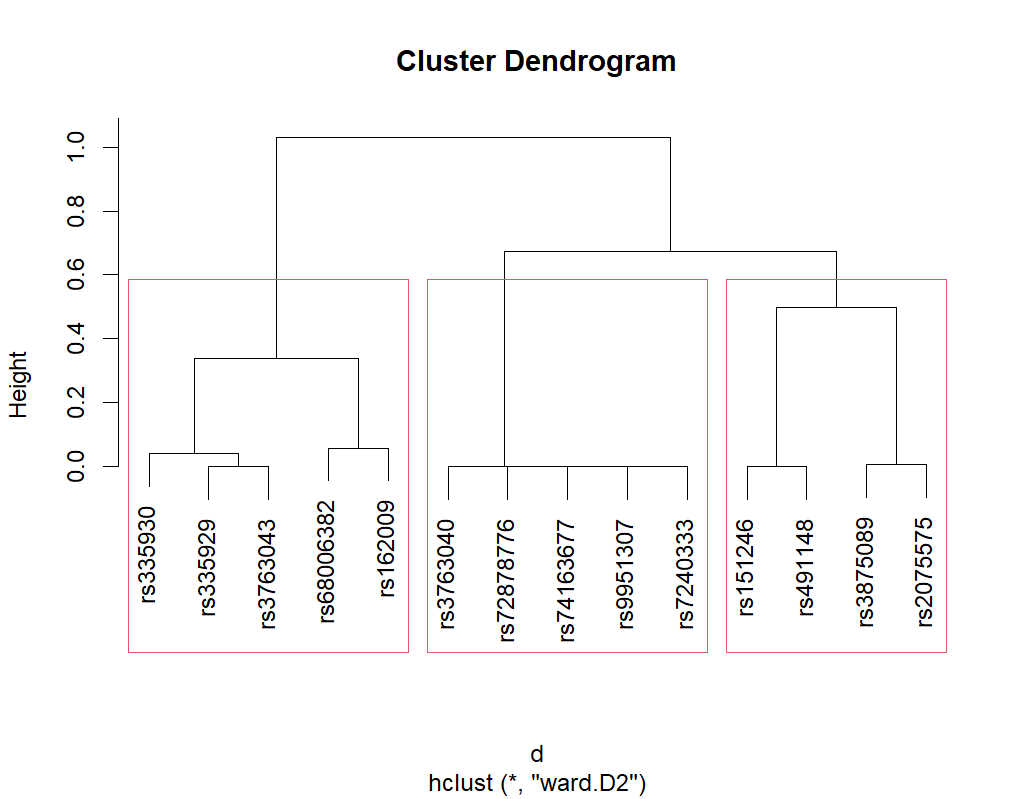

**Figure S2.** Odds Ratios and 95% CI for recessive (0/1 vs 2 minor alleles), dominant (0 vs 1/2 minor alleles) and additive (being carrier of 0, 1 or 2 minor alleles) models in the prediction of presence of PD in *LRRK2* carriers adjusted by age and sex restricted to the G2019S variant carriers subsample (n= 273).

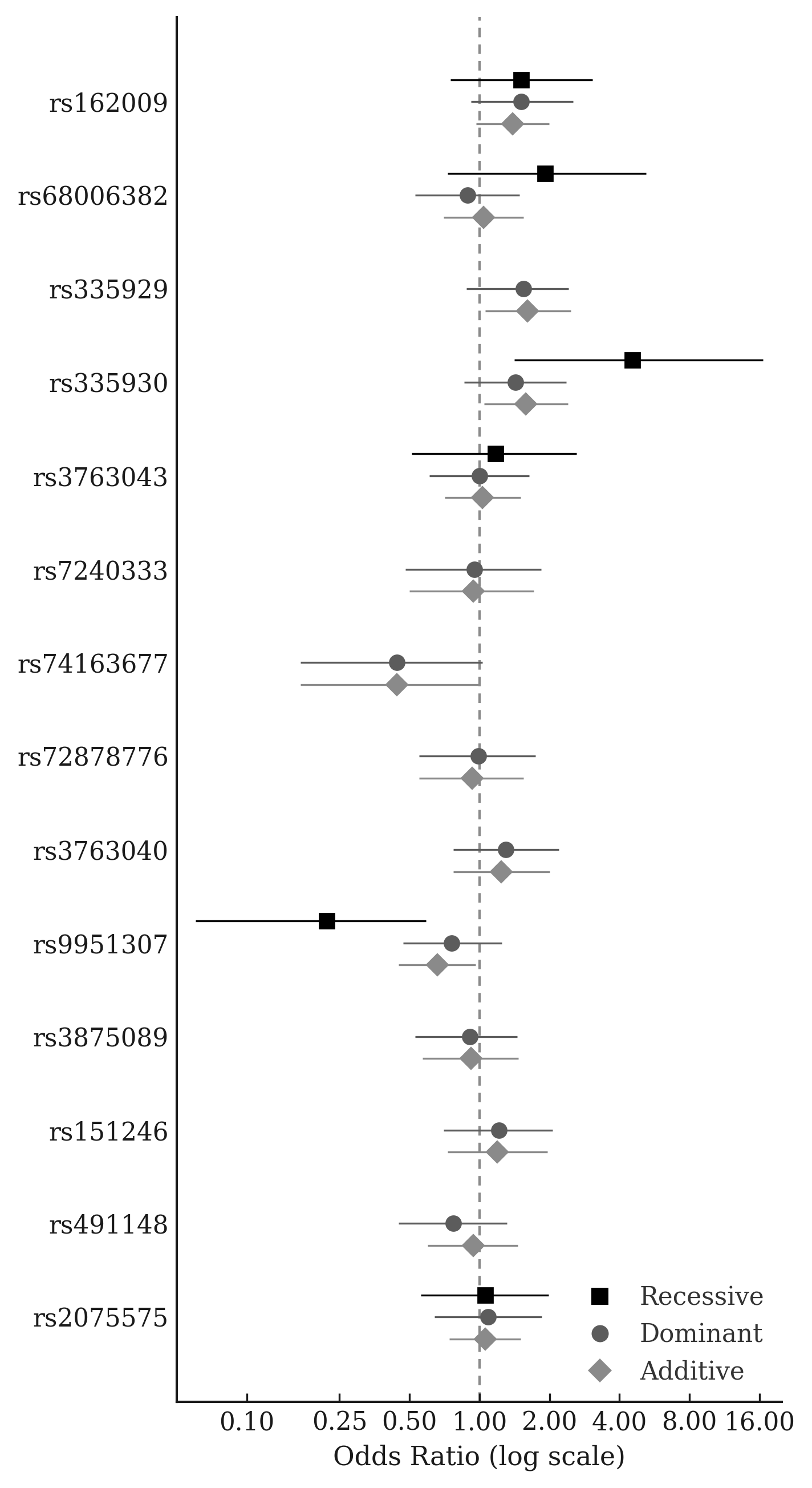
